# Targeted Culture-Independent Sequencing Identifies Emergence of Macrolide-Resistant *Bordetella Pertussis* in Australia

**DOI:** 10.1101/2024.12.19.24319368

**Authors:** Winkie Fong, Rebecca J Rockett, Kingsley King-Gee Tam, Trang Nguyen, Eby M Sim, Enoch Tay, Carl J.E. Suster, Jessica E Agius, Shona Chandra, Anne E Watt, David Speers, Maryza Graham, Thomas Tran, Chuan Kok Lim, Michael C Wehrhahn, Andrew N Ginn, Darcy Gray, Jennifer Robson, Indya Gardner, Rodney McDougall, Lito Papanicolas, Annaleise R Howard-Jones, Alexander C Outhred, Karina Kennedy, Louise Cooley, Qinning Wang, Neisha Jeoffreys, Sharon C-A Chen, Kerri Basile, Tanya Golubchik, Jen Kok, Vitali Sintchenko

## Abstract

*Bordetella pertussis* continues to circulate globally despite wide-spread vaccination, with an emergent international epidemic in 2024. The resurgence of disease is confounded by the emergence of pertactin-deficient, macrolide-resistant *B. pertussis* (MRBP) strains in Asia and Europe, which are under-recognised using traditional diagnostic and surveillance methods. This study addressed these gaps by applying a probe-capture hybridisation technique, which enables targeted culture-independent sequencing of genomes (tNGS) directly from respiratory specimens. Seven co-circulating lineages of *B. pertussis* were identified in Australia, including two associated with MRBP. Eight epidemiologically unrelated and geographically dispersed cases of MRBP in Australia with a A2037G mutation in all three copies of 23S rRNA were documented, three of which were confirmed by phenotypic testing and sequencing of corresponding isolates. The estimated rate of MRBP among *B. pertussis* PCR positive cases was 4.4%. This study demonstrated the value of tNGS based on target enrichment and probe capture sets designed for respiratory pathogens for public health laboratory surveillance of pertussis. This approach can improve the resolution and completeness of *B. pertussis* surveillance given the increasing diversity and vaccine evasion capability of this pathogen.

## Introduction

Pertussis, or whooping cough, is a highly contagious respiratory infection caused by *Bordetella pertussis*, which has been associated with significant morbidity and mortality, especially in young children.^1^ Despite well-resourced vaccination programs in many countries, *B. pertussis* continues to circulate globally, leading to epidemics every 3-5 years.^2^ Following several years of low pertussis incidence during the COVID-19 pandemic, many countries are now experiencing a resurgence of pertussis. In Australia, the incidence of pertussis has substantially increased in 2024 compared to pre-pandemic levels, with over 50,000 cases reported nationally.^3^

This world-wide resurgence of pertussis has coincided with waning immunity among children and adults, increased vaccine hesitancy following the pandemic, and the spread of *B. pertussis* lineages with the potential for vaccine escape due to modification of virulence factors. Acellular pertussis vaccines contain combinations of pertactin, pertussis toxoid, pertussis fimbriae and filamentous haemagglutinin antigens. This induces immune selective pressure at the population level, and supports the spread of pertactin- and/or filamentous haemagglutinin-deficient strains.^4, 5^ In addition, retrospective analyses of historical *B. pertussis* genotyping data suggested a gradual replacement of ancestral strains carrying polymorphism(s) within the pertussis toxin promoter (*ptxP*) from the *ptxP1* allele to newer strains with the *ptxP3* allele, with potentially higher pertussis toxin production.^6^ However, our understanding of emerging pertussis epidemics has become limited due to progressive loss of *B. pertussis* culture capability and over-reliance on nucleic acid amplification tests (NAAT) as the main diagnostic tool. NAAT based assays, especially in a multiplex format, are unable to or cannot provide sufficiently granular data on antigenic profiles to inform vaccination policies or contemporaneous resistance patterns to guide protocolised treatment and prophylaxis approaches.^7^

The emergence of macrolide-resistant *B. pertussis* (MRBP), initially identified in the United States of America in 1993^7, 8^ and subsequently reported in China^9, 10^, has further complicated the control of pertussis epidemics. Alarmingly, MRBP has spread globally with cases recently reported in China^10^, Vietnam^11^, Japan^12, 13^ and Europe.^14^ Some evidence suggested that MRBP was initially associated with the *ptxP1* genotype but lately MRBP populations appear to be dominated by strains containing the potentially more virulent *ptxP3* genotype.^15^ Macrolides, specifically erythromycin and azithromycin, have been first-line treatment options for pertussis cases and their contacts; clinical data on effectiveness of second line agents, such as trimethoprim/sulfamethoxazole, are lacking. These changes in molecular epidemiology and the increase in MRBP highlight key deficiencies in current public health laboratory surveillance strategies for pertussis and the role of prospectively genotyping *B. pertussis* strains.

This study aims to address these gaps by applying a targeted whole-genome sequencing by probe-capture hybridisation, which specifically enriches *B. pertussis* from clinical specimens, and enables full-genome culture-independent sequencing directly from specimens. The targeted next generation sequencing (tNGS) analysis was benchmarked against culture-based sequencing of a subset of RT-PCR *B. pertussis* as well as culture positive specimens in our cohort. The described approach not only facilitates the detection of macrolide resistance mutations, but provided a comprehensive picture of circulating *B. pertussis* populations co-circulating in Australia in 2024 that is relevant for public health interventions and treatment guidelines.

## Materials and Methods

### Clinical specimens

Respiratory specimens (n = 255) where *Bordetella* DNA was detected by RT-PCR targeting IS*481* with cycle threshold (Ct) <30 was included in the study. This cohort included nasopharyngeal aspirates (n = 17), sputum (n = 1), nasopharyngeal/throat swabs (n = 212), and unknown respiratory specimen types (n = 25) collected between January and October 2024 and tested by the participating laboratories. They represented all states and territories of Australia with the exception of the Northern Territory, and included New South Wales (NSW): NSW Health Pathology-Institute of Clinical Pathology and Medical Research (ICPMR), The Children’s Hospital at Westmead and Douglass Hanly Moir Pathology; Queensland (QLD): Sullivan Nicolaides Pathology; South Australia (SA): SA Pathology; Victoria (VIC): Victorian Infectious Diseases Reference Laboratory; Western Australia (WA): PathWest Laboratory Medicine; Australian Capital Territory (ACT): ACT Pathology; Tasmania (TAS): Royal Hobart Hospital.

### B. pertussis-specific PCR

All specimens included in the study were confirmed as *B. pertussis by* an in-house PCR that differentiates between *B. pertussis* (presence of IS*481*)*, B. parapertussis (*IS*1002)* and *B. holmesii (*hIS*1001)*.^16^

### Macrolide-resistant B. pertussis (MRBP) PCR

All specimens included in the study were screened for MRBP by PCR described using the primers and probes listed in Table 1. The PCR mastermix utilised 500nM of each primer, 100nM of each probe and Immomix 2X buffer (Bioline). The total reaction volume comprised of 10 μL of template DNA, and 15μL of mastermix. A thermocycling profile of 95 C for 10 minutes, followed by 45 cycles of amplification at 95 C for 10 s, 48 C for 30 s, 72 C for 30s, performed on a LightCycler 480 II (Roche Life Science) and a colour compensation was applied during analysis.

**Table 1:**
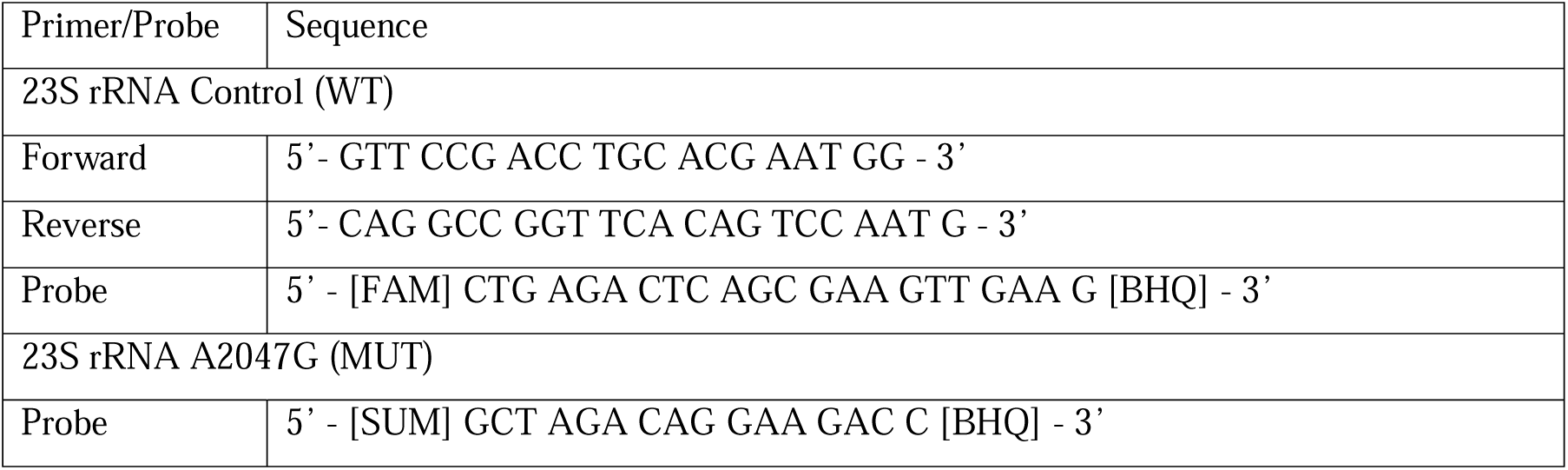
Primers and probes utilised in the macrolide-resistant *B. pertussis* (MRBP) PCR.

### Targeted next generation sequencing (tNGS) via probe-capture

*B. pertussis* closed reference genomes (n = 18, Supplementary Material 1) were used to develop a custom probe panel which tiled the complete *B. pertussis* genome x1 using CATCH.^17^ Probes were synthesised by Twist Biosciences. Genomic DNA from clinical specimens tested at ICPMR and externally referred clinical specimens were extracted using NucliSENS easyMag (bioMérieux). Other participating laboratories submitted a mixture of DNA extracts used in their PCR testing from either the MagNA Pure (Roche Life Science) or QIAamp 96 Virus QIAcube HT Kit on the QIAcube HT System (Qiagen) and/or residual clinical specimens. Libraries were prepared from genomic DNA with the Twist EF (version 1) library preparation kit with protocol miniaturisation to a quarter of manufacturers volume. Up to 96 libraries were pooled in equal volumes and vacuum concentrated to a dry pellet before being hybridised for 17 hours using Twist standard hybridisation reagents and custom probes. Enriched libraries were then sequenced on the NextSeq 2000 platform (Illumina) using P1 or P2 kits with the aim of generating a minimum of 3 million reads per library.

### Bioinformatic analysis of tNGS data

The sequenced raw reads were first subjected to an in-house quality control procedure consisting of fastp (version 0.22.0)^18^, PRINSEQ (v.1.2.4)^19^ and Kraken2 (v.2.0.8).^20^ The sequencing data were then mapped with Minimap2 (v.2.22-r1101)^21^ to *B. pertussis* H640 (CP025371.1)^22^, *B. holmesii* (CP043146.1)^23^ or *B. parapertussis* (CP025070.1)^23^ genomes as references. The reference genome was selected depending on the PCR result of each specimen. The genome coverage and depth of sequencing were evaluated using Samtools (v.1.11).^24^ The reference genome was selected depending on the PCR result of each specimen. Human DNA reads were then depleted from quality checked target bacterial reads through a combination of Kraken2 identification using a custom human pangenome database^25^ and Bowtie2 mapping to the GRCh38.p13 genome.^26^ The remaining reads were then passed through ‘pertpipe’ (https://github.com/cidm-ph/pertpipe) using the --meta flag. This approach utilised assemblies generated by metaSPAdes (v.3.15.5)^27^ for virulence gene detection, and both MEGAHIT (v.1.2.9)^28^ and kallisto (v.0.46.0)^29^ for identification of mutations conferring antibiotic resistance. Briefly, the MEGAHIT assembly was aligned with a single copy of the 23S rRNA from *B. pertussis* strain Tohama I (NC_0029292.2), and reference mapped with kallisto to the SILVA ribosomal RNA gene database.^30^ Kallisto reference mapping --pseudobam was performed on the human-depleted reads using a custom database of genomes (Supplementary Material 1). The mapped *B. pertussis* reads were then extracted and uploaded to BioProject: PRJNA1199062. These reads were assembled with standard SPAdes. Genomic relatedness between kallisto-filtered tNGS and WGS assemblies were assessed via a *B. pertussis* core genome multi locus sequence typing (cgMLST) schema.^31^ Briefly, assemblies were filtered by SeqKit (v 2.4.0)^32^ to capture all contigs greater than 300-bp and a *B. pertussis* training file was generated for the genome of *B. pertussis* strain Tohama I (NC_002929.2) using Prodigal (v 2.6.3).^33^ The *B pertussis* schema was adapted and run using chewBBACA (v 3.3.10).^34^ Conversion of cgMLST results into a distance matrix was performed using cgMLST-dist (https://github.com/tseemann/cgmlst-dists). The human-depleted reads were analysed through an in-house metagenomics pipeline for identification of potential co-infections (https://github.com/YasirKusay/meta_transcriptomics_pipeline_sample).

### Isolation of B. pertussis

Clinical isolates (n = 45) were recovered on Charcoal Blood Agar (CBA) supplemented with 40 μg/mL cephalexin from corresponding respiratory aspirates or swabs by reflex culture conducted at ICPMR (n = 14), Douglass Hanly Moir (n = 17), The Children’s Hospital at Westmead (n = 5) and Sullivan Nicolaides Pathology (n = 9) laboratories. Isolates were collected between 2015-2019 (n = 5) and in 2024 (n = 40). Minimum inhibitory concentrations (MICs) for erythromycin and azithromycin were determined by Etest (bioMérieux, France) on CBA plates inoculated with bacterial suspensions of 0.5 McFarland in saline. All plates were incubated for 48 hours at 37 C under aerobic conditions.

### Whole genome sequencing of B. pertussis isolates

Whole genome sequencing (WGS) was performed at the Centre for Infectious Diseases and Microbiology-Public Health (CIDM-PH), Westmead Hospital. Genomic DNA was extracted from cultures with either the DNeasy Bloodand Tissue Kit or the DNeasy UltraClean Kit (Qiagen). Sequencing libraries were prepared using either the Nextera XT DNA Library Prep Kit (Illumina) or Nextera DNA Prep Kit (Illumina) as per the manufacturers’ instructions and sequenced on a NextSeq 500 (Illumina) using NextSeq 500/550 V1 or V2 kits (Illumina), or NextSeq 2000 using P1 or P2 kits. Where required (n = 4), the same DNA extracts were employed as input for long-read Nanopore sequencing (Oxford Nanopore Technologies, ONT). Libraries were prepared using the SQK-RBK004 rapid barcoding kit (ONT) according to manufacturers’ instructions and loaded onto a R.9.4.1 flow cell. Sequencing was performed on a MinION Mk1B running MinKNOW v.24.02.16 with live base calling on high-accuracy mode.

### WGS bioinformatics

The raw reads from culture-based WGS were checked for quality through an in-house pipeline consisting of FastQC (v.0.11.3), Trimmomatic (v.0.36)^35^ and Centrifuge (v.1.0.4)^36^, prior to further analysis. Quality checked reads were then analysed with pertpipe in its genomic DNA mode. This mode employed SPAdes (v.3.15.5)^37^ for assembly of reads and utilised a combination of BLAST+ (v.2.9.0),^38^ Minimap2 (v.2.22-r1101),^21^ MLST (v.2.22.1; https://github.com/tseemann/mlst)^39^ and ABRicate (v.0.9.8; https://github.com/tseemann/abricate)^40^ to identify alleles of *ptxP, ptxA-E, prn, fhaB, fim2* and *fim3*, as well as the 23S rRNA, which were confirmed through both assembly and mapping. All *B. pertussis* genomes shared by Weigand *et al*^22^ (BioProject: PRJNA279196), Xu *et al*^41^ (BioProject: PRJNA489102) and Cai *et al*^10^ (BioProject: PRJNA908268) were included and examined for genomic context. A further set of closed genomes was downloaded from National Center for Biotechnology Information Reference Sequence Database (NCBI RefSeq), selecting a randomly selected subset with the USA as the country of origin to limit over representation from a high number of sequences from a single country. A core gene alignment was produced with Panaroo (v.1.2.7) and a phylogeny was inferred with IQ-TREE (v.1.6.7) using the GTR+F+R2 model, which was selected by ModelFinder. A core single nucleotide polymorphism (SNP) analysis was performed with Snippy, Snippy-core (v.4.6.0) and SNP-dists (v.0.6) and a phylogeny was inferred with IQ-TREE using the GTR+F+R2 model. Phylogeny and bioinformatic analysis were visualised using R studio with the packages ggtree^42^, ggplot2^43^, and BactDating (https://github.com/xavierdidelot/BactDating).

ONT long-reads were filtered using Filtlong (v.0.2.1; https://github.com/rrwick/Filtlong) to include only reads greater than 1000 base pairs in length. Centrifuge (v.1.0.4)^36^ was used to assess for contamination of ONT outputs. Assembly and polishing, using both short-and-long reads was performed using Dragonflye (v.1.2.1; https://github.com/rpetit3/dragonflye). The long-read assembler chosen was Flye (v. 2.9.5-b1801)^44^ using the following flags “-g 4100000 --asm_coverage 100”. One round of long read polishing was performed using the “r941_e81_hac_g514” model on Medaka (v.1.11.3; https://github.com/nanoporetech/medaka) and one round of short-read polishing was performed using Polypolish (v.0.6.0)^45^

## Results

### Recovery of Bordetella spp. genomes from clinical specimens by tNGS

Within the study, 255 clinical specimens were subjected to tNGS and *B. pertussis* genomes were recovered in 146 specimens using this approach (57.2%). Fourteen specimens (or 5.5%) were found to contain either *B. holmesii* (n = 8) or *B. parapertussis* genomes (n = 6) (**Figure 1**). A low genome recovery rate of these alternative causes of pertussis disease via this method was observed. Overall, the recovery of *B. parapertussis* and *B. holmesii* ranged between 0.53-69.12% genome coverage, and 0.007-14.97 average depth. No instances of co-infections between the various pertussis causing organisms were observed. All successfully recovered *B. pertussis* genomes (n = 146) were associated with greater than 98% genome coverage and over 10X average depth (**Figure 2A**), with an average of 11 million total reads (range 679,000 to 109 million reads) per target genome. The depth of sequencing was evenly distributed across the genome (**Figure 2B**). Successful recovery of *B. pertussis* from tNGS correlated with initial Ct values from the diagnostic RT-PCR performed at ICPMR for routine microbiology, where the highest Ct value with a successful sequence was Ct 28. However, a significantly lower sequencing success rate was observed in specimens with PCR Ct values greater than 25 (Supplementary Figure S1). High abundance of human cells and commensal bacterial flora in respiratory samples negatively affected the retrieval of *B. pertussis* genomes, and where the capture efficiency of *B. pertussis* was less than 50%, consistent recovery of *Bordetella* genomes became difficult (Supplementary Figure S1 & S2).

**Figure 1.**
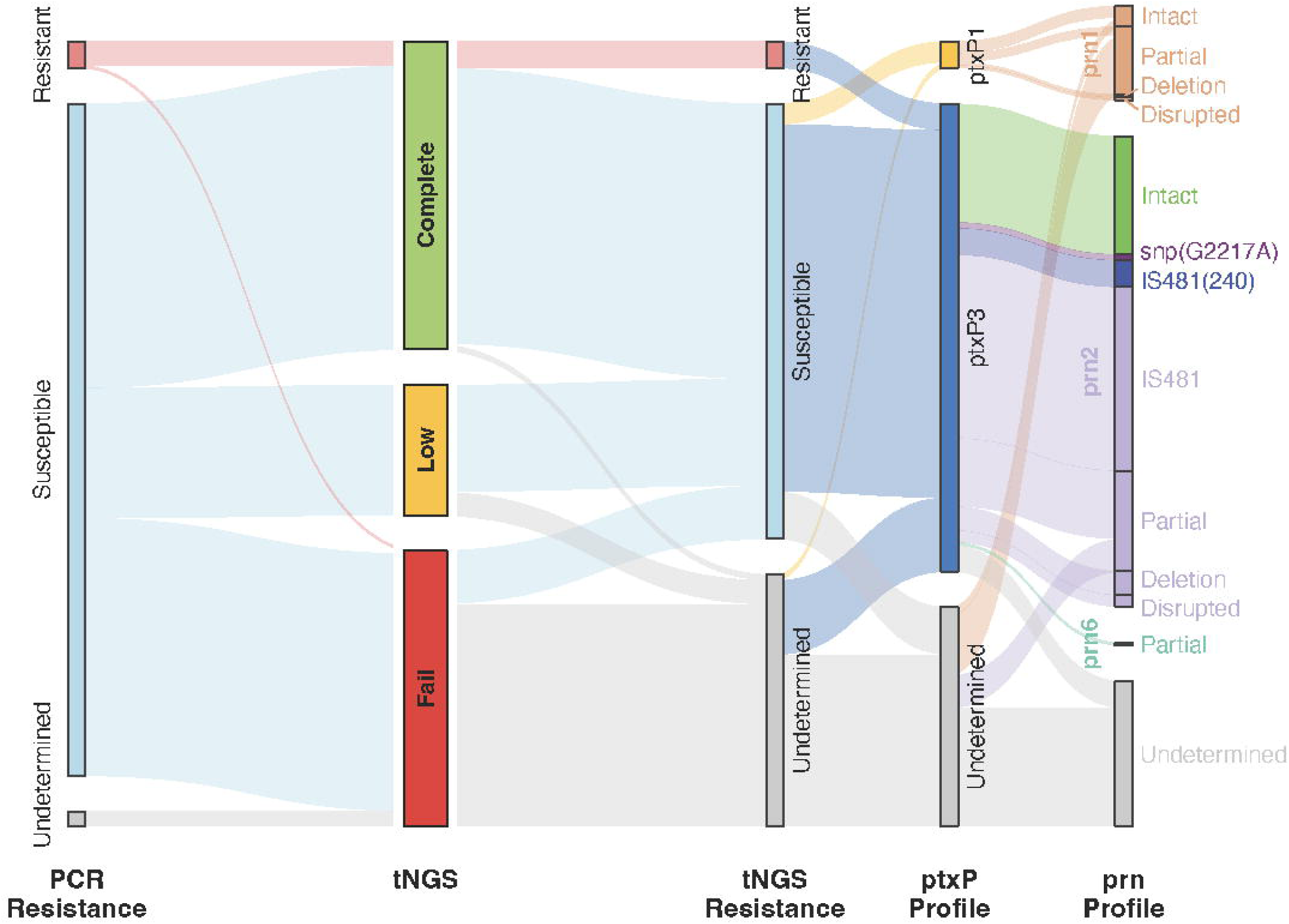
Sankey chart distributing all 255 specimens attempted within the study, including genomic features pertussis toxoid promoter (*ptxP*) type and pertactin (*prn*) type. Partial typing of *prn* genes was included from Low-Fail specimens however a partial *prn* assignment does not indicate a disruption of *prn* potentially resulting in *prn-*negative expression.

**Figure 2.**
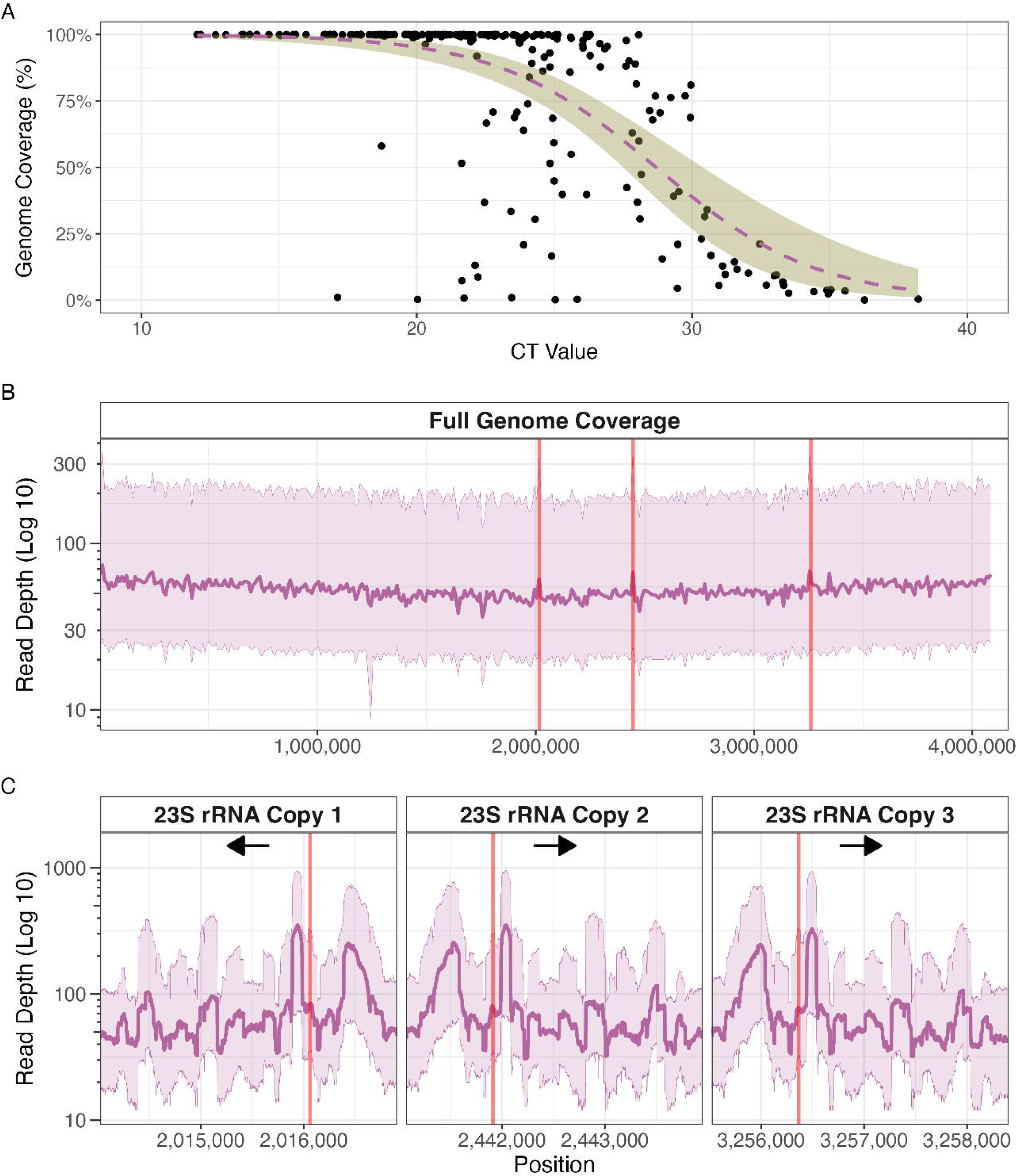
Retrieval of informative *B. pertussis* genome sequences from diagnostic respiratory samples. **(A)** Scatterplot of percentage of genome coverage and CT value, with the purple dashed representing the binomial GLM line, and olive shading representing the 95% confidence interval. Ct values more than 25 started observing a sharper decline in genome recovery. **(B & C)** The coverage depth of all successful genomes (n = 146) over the whole genome and 23S rRNA region. The dark purple line is the median coverage, flanked by the lighter purple lines as the 25% and 75% interquartile ranges. The locations of three copies of 23S RNA sequences in the whole genome and the A2037G mutation conferring resistance to macrolides within the 23S rRNA are indicated by red lines, arrows represent the positive and negative strand (orientation) of the 23S rRNA in the reference.

### Diversity of B. pertussis populations in 2024 snapshot

One hundred and seven out of 183 (58.4%) clinical specimens were successfully sequenced with adequate genome coverage for macrolide resistance analysis. Of the 107 specimens, 96 yielded sufficient data to infer complete virulence typing information (*ptxP*, *ptxA*, *prn*, *fim3* and *fhaB*). Seven distinct virulence profiles were observed within our dataset from the 87 tNGS sequences, and nine culture-derived WGS sequences (**Figure 3**). The predominant virulence profile was *ptxP3/ptxA1/prn2*(neg)/*fim3-2/fhaB1* (n=43, 44.8%), followed by *ptxP3/ptxA1/prn2*(pos)*/fim3-1/fhaB1* (n=20, 20.8%). For each individual virulence allele within the virulence profile, the analysis successfully determined *ptxP* types from 123 sequences, with 116 *ptxP3* (94.3%) and 7 *ptxP1* (5.7%). In the case of *ptxA* alleles, 113 sequences were analysed, and they were all identified as *ptxA1*. Co-circulating *B. pertussis* strains more commonly carried *prn2* (n=119, 80.4%) than *prn1* (n=29, 19.6%). The pertpipe analysis of these *prn* types in 101 sequences further classified *prn1* and *prn2* subtypes into *prn 1* intact (n=5, 5.0%)*, prn2* intact (n=32, 31.7%) and subtypes with different genomic evidence of pertactin allelic disruption. Specifically, we documented *prn1*::del(1757, 2379) (n=1, 0.9%), *prn1*::dis(2475) (n=1, 0.9%), *prn2*::del(−292, 1340) (n=2, 1.9%), *prn*2::del(1080, 1345) (n=1, 0.9%), *prn2*::del(1984, 1986) (n=1, 0.9%), *prn2*::dis(2748) (n=4, 4.0%), *prn2*::IS*481*(1613) (n=44, 43.6%), *prn2*::IS*481*(240) (n=8, 7.9%), and *prn2*::snp(G2217A) (n=2, 1.9%). For the breakdown of *fim3*, 75 sequences classified as either *fim3-1* (n=28, 37.3%), *fim3-2* (n=35, 46.7%) or *fim3-26* (n=12, 16.0%). In *fhaB,* truncation of the gene as a result of non-contiguous assembly occurred in 111 of 150 sequences (70.3%), and in these instances *fhaB* type was determined using only the longest contig, which could result in misassignment of allele type. From these 150 *fhaB* genes, 109 were *fhaB1* (truncated in n=70, 64.2%), one was *fhaB*2 (truncated in n=1, 100%) and 40 were *fhaB3* (truncated in n=40, 100%).

**Figure 3:**
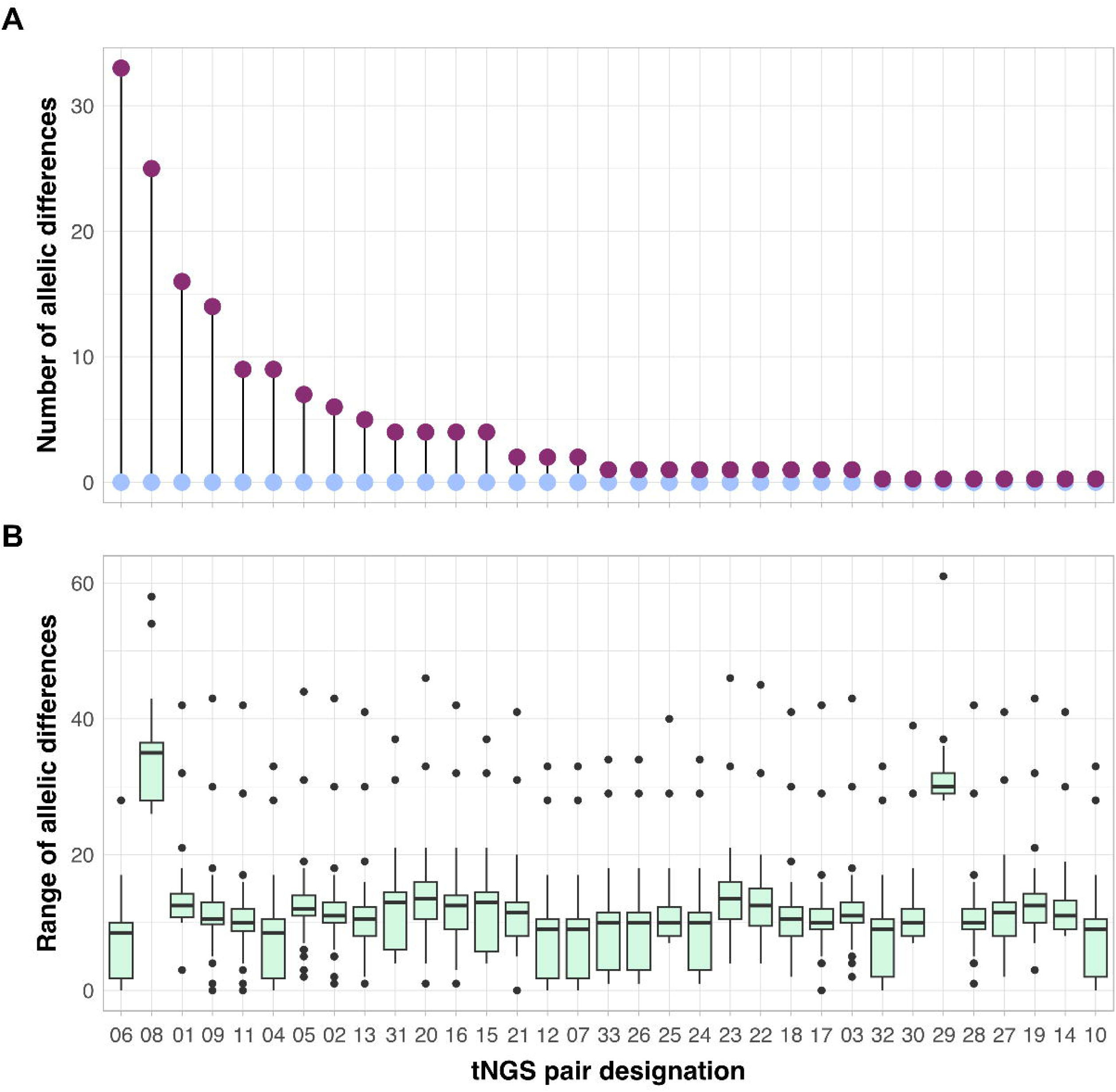
Assessment of genomic concordance between the 33 paired targeted next generation stNGS and whole genome sequencing (WGS) *B. pertussis* assemblies via core-genome multi-locus sequence typing (cgMLST). **(A)** Number of allelic differences between each paired tNGS and WGS sample. **(B)** Range of allelic differences between each tNGS assembly to all other non-paired WGS assemblies.

### Concordance of sequencing data derived from tNGS and WGS

There were 45 clinical specimens with a paired *B. pertussis* isolate, which were assessed for concordance to determine the accuracy of probe-capture derived data compared to WGS. Of the 45 pairs, 33 were sequenced successfully with both tNGS and WGS, and of these 20 pairs shared the same virulence profile and had fewer than 5 core genome SNPs difference with their paired sequence. Genomes with different degrees of discordance (13 pairs) appeared to have slightly larger core distances i.e. over 5 SNPs (n=10), discordant *fim3* (n=1)), discordant (n=8) subtypes, or a combination of above (**Figure 3**).

### Emergence of MRBP in Australia

Screening of the 23S rRNA sequence from probe capture sequencing and the MRBP PCR revealed that ten specimens collected from eight individual cases contained the A2037G mutation conferring resistance to macrolides. These eight MRBP cases were from NSW (n=6), Victoria (n=1), and Queensland (n=1). Of the MRBP cases, seven were from children (<18 years old) and one from an adult. All eight cases were classified into two distinct virulence profiles *ptxP3/ptxA1/prn2*(neg)/*fim3-1/fhaB1* (n=6) and *ptxP3/ptxA1/prn2*(pos)*/fim3-1/fhaB1* (n=2) (Supplementary Figure S3), and further *prn* gene analysis revealed them to be *prn2*::*IS481*(240), which is *prn2* negative, and *prn2*::snp(G2217A), which is an intact *prn2*. The allelic frequency of A2037G mutation 23S rRNA in these eight specimens ranged from 91.7-100%, suggesting that the macrolide-conferring resistant mutation was likely present in all three copies of the 23S rRNA.

From three of eight cases (four of ten specimens), MRBP was successfully cultured from their corresponding respiratory specimens. Phenotypic susceptibility testing confirmed macrolide resistance in all four isolates (one case with two isolates) with an MIC of >256 mg/L for both erythromycin and azithromycin. Through hybrid assembly, we confirmed that each of the four genomes had the A2037G mutation in all three copies of their respective 23S rRNA. Further, the application of the MRBP PCR on all specimens detected the resistance conferring mutation in 9/10 specimens, the phenotypically resistant case with a susceptible PCR result was the case with the 91.7% allelic frequency.

Most MRBP genomes in this study were genomically distinct, and there were no apparent epidemiological links between the cases in NSW. Comparison of our MRBP sequences to those reported from Shanghai, China in 2021-2022, revealed similarities between newly detected Australian MRBP and those from China (**Figure 4A**). SNP distances between MRBP isolates from Australia and China ranged from 1 to 79 core SNPs, with a median of 16 core SNPs. Bayesian phylogeny of MRBP strains suggest some of the strains detected in Australia may have recently spread from China (**Figure 4B**).

**Figure 4:**
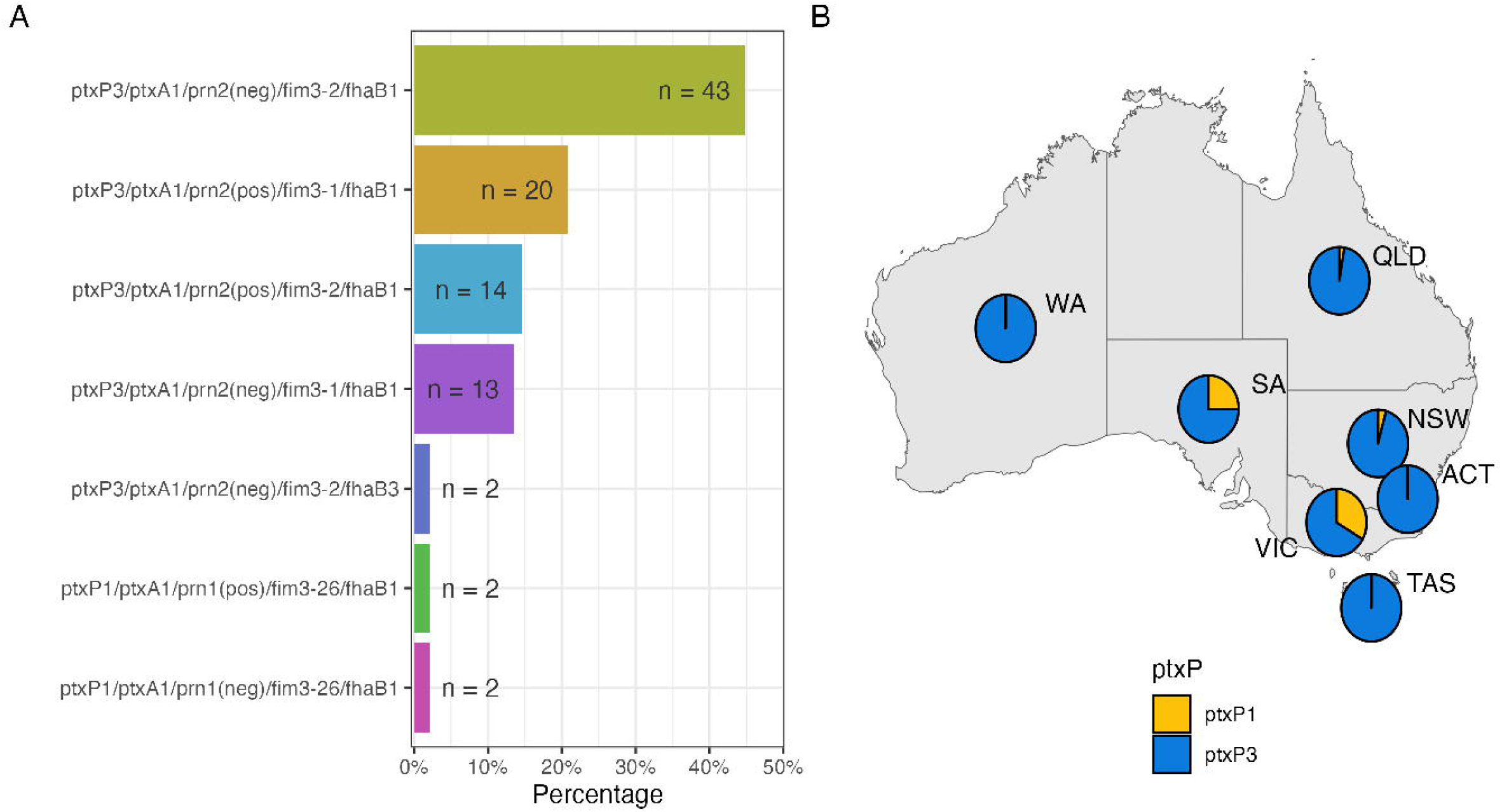
Genomic and geographical diversity of *B. pertussis* genomes in the 2024 snapshot. **(A)** Distribution of virulence profiles (n = 96) observed during the 2024 *B. pertussis* outbreak. **(B)** Distribution of sequences with pertussis toxoid promotor (*ptxP*) information across Australia during the 2024 snapshot period (n = 123). State numbers as follows Australian Capital Territory (ACT) (n = 2), New South Wales (NSW) (n = 69), Queensland (QLD) (n = 37), South Australia (SA) (n = 8), Tasmania (TAS) (n = 1), Victoria (VIC) (n = 3), Western Australia (WA) (n = 3). Virulence profiles and *ptxP* alleles were determined from targeted next generation sequencing (tNGS) sequences, and if tNGS virulence profiles from paired specimens were incomplete or missing, then the WGS virulence profiles were supplemented.

**Figure 5:**
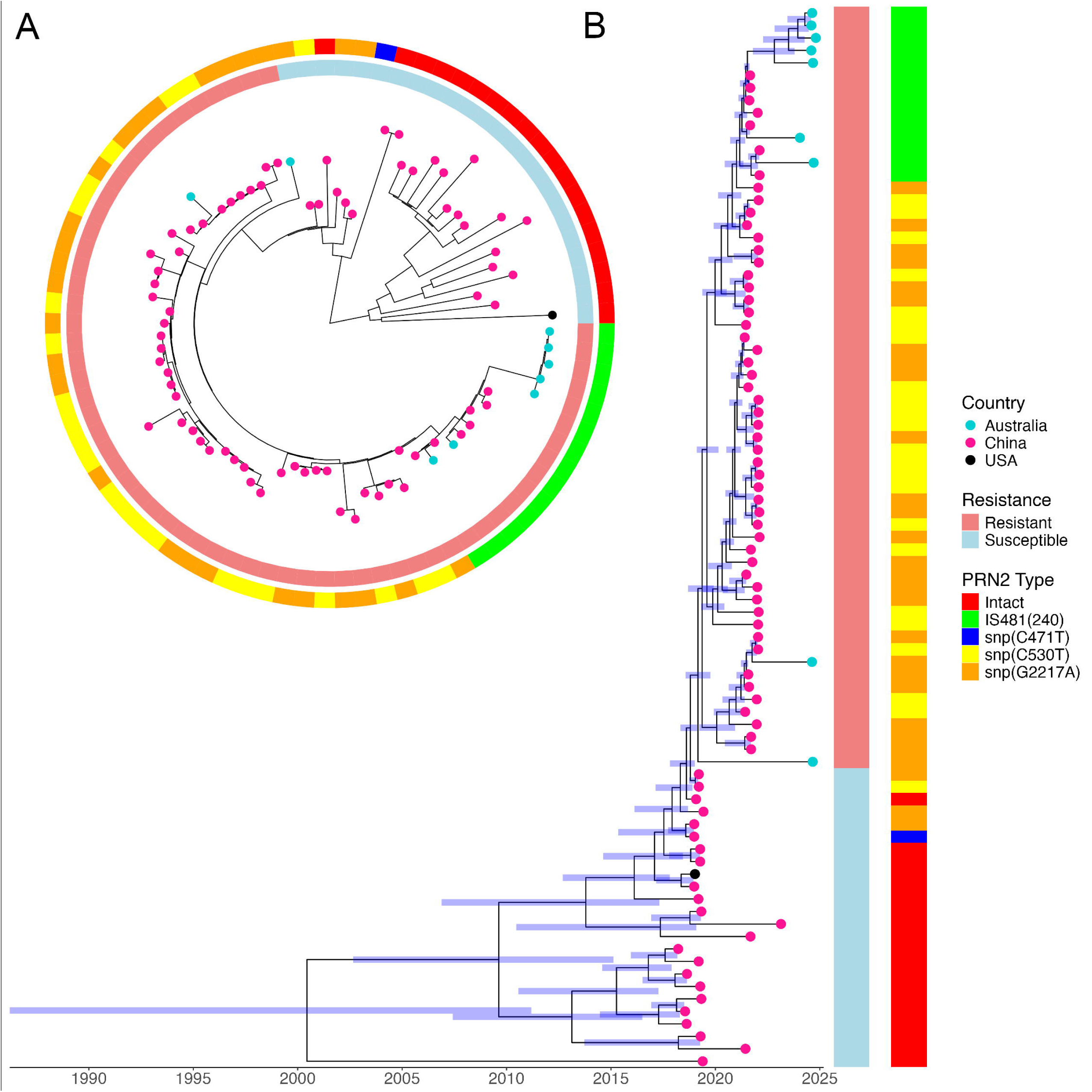
Relationships between MRPB genomes from cultured isolates in Australia and the ancestral *B. pertussis* genomes. **(A)** Core single nucleotide polymorphisms (SNP) phylogeny of the Australian macrolide-resistant *B. pertussis* MRBP genomes (n = 9, from 7 cases) from targeted next generation sequencing (tNGS) in comparison to all recent MRBP detected in Shanghai, China (Bioproject: PRJNA908268), the black dot tip represents the Reference genome H640 (CP025371.1). **(B)** Bayesian phylogenetic inference of recent ancestors of Australian strains of MRBP in the international context.

## Discussion

This study adds important insights to the current epidemic of pertussis affecting countries world-wide. Our findings reconfirmed the presence of pertactin-deficient variants in Australia, a country with high childhood acellular pertussis vaccine coverage rates following its introduction into the National Immunisation Program in 1996 (replacing the whole cell vaccine).^6^ This study provided a nationally representative and high-resolution snapshot of *B. pertussis* diversity which is largely free from the sampling bias introduced by bacterial culture, from which typically only a single dominant strain from a diagnostic specimen is characterised. We documented co-circulation of seven distinct virulence profiles. Most of the specimens interrogated carried both the *ptxP3* and disrupted pertactin gene allele, which is concerning as pertactin-deficient *B. pertussis* strains appear to exhibit increased fitness within vaccinated populations^5^ and previous studies have determined *ptxP3* strains are more virulent in humans than *ptxP1*, as *ptxP3* strains produce almost double the amount of pertussis toxin.^46^

Culture-independent probe capture tNGS successfully identified cases of MRBP and estimated the incidence of MRBP in our Australian cohort at 4.4% in 2024. At least half of these detections were verified by corresponding culture results. The one discordant MRBP PCR and tNGS result could be explained by a subpopulation of susceptible *B. pertussis* within the sample, as not all reads detected in tNGS carried the A2037G mutation, therefore potentially increasing the Ct value beyond detectable limits. As the *IS481* PCR Ct value was 24.51, in contrast to 32.99 in the control target in the MRBP PCR, a subpopulation would have influenced the PCR. Additionally, MRBP Case #8 failed tNGS QC cut-offs, and was identified by the MRBP PCR, however virulence profile information was still able to be determined and the 23S rRNA A2037G mutation was supported by 5 total reads (below the 10 read cut-off). Notably, the cohort of MRBP cases in this study was represented by two virulence profiles, suggesting co-circulation of MRBP strains from independent incursions into Australia. No evidence of treatment-induced MRBP was established in this study, as detections of the A2037G mutation were mainly from specimens that were collected prior to treatment. Analysis of core genome SNP phylogeny indicated genetic similarity of Australian MRBP strains to those recently reported from China, albeit without epidemiological evidence of direct importation from China to Australia. Of note, the lack of available MRBP sequences from other countries where MRBP was reported prevented accurate determination of potential origins of Australian MRBP. Given the clonality of *B. pertussis* populations and a mutation rate of 0.9 SNPs/per year (2.24×10^−7^ per site/year)^47^, it is important to establish and maintain genomic surveillance with sufficient coverage over virulence alleles such as *ptxP3* and 23S rRNA to monitor the international spread of MRBP. In addition, such a genomic surveillance strategy allows detection of other mechanisms of resistance that may emerge in the future.

The sensitivity of the presented approach is dependent on bacterial load of the target microorganism, thus we only attempted culture-independent sequencing on specimens with *B. pertussis* RT-PCR Ct value ≤25. Probe capture tNGS recovered *B. pertussis* genomes with over 98% coverage and over 30-fold average depth. However, genomes with less than 30-fold average depth were more difficult to assemble with confidence, resulting in a higher likelihood of abnormal truncation of genes, particularly in the 23S rRNA and the *prn* gene regions. Without accurate data from these genes, assessment of the virulence profile and macrolide resistance detection remains problematic. Often, despite low Ct values of some clinical specimens and target enrichment, it is suspected that contamination with human or commensal bacteria DNA contamination still significantly affected the recovery of *B. pertussis* genomes. To prove this theory shotgun metagenomics could be performed on the same clinical specimens.

Our findings demonstrated the high specificity of probe-capture tNGS approaches. We have filtered out 23S rRNA sequences present in bacterial species other than *B. pertussis* using SILVA database in order to minimise the risk of mis-mapping the 23S rRNA from other respiratory flora. While ample care was taken to remove noise from other species, other mutations in the 23S rRNA were occasionally observed, albeit at a significantly lower frequency, which did not affect consensus 23S rRNA sequences (∼3.2%). There was minimal cross-reactivity of the probes between *B. pertussis, B. parapertussis* and *B. holmesii*. However, the high Ct values and correspondingly low abundance of other *Bordetella* spp. in the study set may have influenced the low cross-reactivity observed.

We acknowledge other potential limitations of the study. First, the success of tNGS methodology depends on the yield of the target pathogen in a clinical specimen and it is likely that more severe pertussis cases with a higher load of *B. pertussis* are over-represented in our cohort. Second, the specimens provided by partner laboratories to this snapshot represented only a small fraction (less than 1%) of all cases of laboratory-confirmed pertussis in Australia in the study period. Nevertheless, the broad representation of community and hospital based clinical specimens from different jurisdictions, the relatively high number of specimens in comparison to other pertussis epidemiology studies and high-resolution of culture-independent sequencing support the quality of data and study inferences. The additional and unique strength of this study was the availability of *B. pertussis* cultures paired with a subset of clinical specimens subjected to tNGS for comparison. The concordance of *B. pertussis* virulence profiles identified by culture-independent tNGS on clinical specimens and from the genomes of cultured isolates from the same specimens provided an additional layer of validation. A total of 13 pairs were somewhat discordant between tNGS and WGS, a result of subpopulation of alternative strains within the clinical specimen, cross-mapping of reads from other closely related species, or low coverage across certain regions resulting in poor assemblies and therefore potentially inaccurate calls of *prn* types or *fim3* types. Additionally, the utilisation of metagenomic assembly tools could have contributed to these discrepancies.

In conclusion, tNGS using hybridisation probes designed to enrich the complete *B. pertussis* genome identified distinct virulence profiles of *B. pertussis* strains co-circulating in Australia in 2024 with an estimated MRBP rate of 4.4%. This highlights the need for enhanced surveillance for MRBP nationally and internationally and necessitates re-evaluation of current microbiology testing practices for pertussis. Targeted sequencing enables culture-independent recovery of almost complete *B. pertussis* genomes directly from clinical specimens with sufficient yield of the target pathogen. Our findings address the increasingly recognised need to improve the completeness and resolution of surveillance for *B. pertussis* given the growing diversity and vaccine evasion capabilities of this pathogen. The introduction of tNGS offers the ability to sequence fastidious bacteria such as *B. pertussis* directly from respiratory specimens without significant delays as this method requires only a single overnight capture on a PCR machine prior to sequencing, which provides timely public health surveillance for control of pertussis and other respiratory pathogens.

## Acknowledgements

The authors would like to thank the Microbial Genomics Reference Laboratory and the Centre for Infectious Diseases and Microbiology Laboratory Services for their expertise and routine sequencing of *Bordetella* spp. genomes utilised in this study. We are indebted to diagnostic laboratories for referring *B. pertussis*, *B. holmesii* and *B. parapertussis* specimens to our centre for typing and characterisation. The authors acknowledge the Sydney Informatics Hub, a Core Research Facility of the University of Sydney and their high-performance computing cluster Artemis for providing the compute capabilities to analyse these genomes.

## Data Availability

Genome sequences generated in the study were uploaded to NCBI SRA and GenBank Database under BioProject: PRJNA1199062 and PRJNA1178746. Individual reads and assembly accessions are provided in Supplementary File S2.

## Competing Interests

All authors declared no conflict of interest.

## Author Contributions

W.F designed and developed the Python pipeline and drafted the manuscript. The study was conceptualised by W.F., R.R., J.K, and V.S. Specimens were supplied by D.S., M.G., T.T., C.K.L., M.W., A.G., D.G., J.R., I.G., R.M., L.P., A.H.J., A.O., K.K., L.C., and N.J., Isolate collection, identification and subculturing was performed by T.N. Clinical specimen processing and collection was managed by W.F., J.A., and S.C. Clinical specimens were extracted by N.J., and her staff at the Centre for Infectious Diseases and Microbiology Laboratory Services, NSW Health Pathology-Institute of Clinical Pathology and Medical Research. Targeted metagenomic sequencing of specimens were conducted by K.T. and R.R. The MRBP PCR was developed by E.T., and performed by E.T., and K.T. Culture-dependent WGS was performed by Q.W., K.B., and the staff at the Microbial Genomics Reference Laboratory. Nanopore sequencing was performed by E.S., Bioinformatics and genomic epidemiology was performed by W.F., E.S. and A.W. Figures were generated by W.F. and C.S. All authors read and approved the final manuscript.

## Ethics Declaration

Genomes and metadata were collected by the Microbial Genomics Reference Laboratory at the NSW Health Pathology-Institute of Clinical Pathology and Medical Research under the Western Sydney Local Health District Human Research Ethics and Governance Committee (Project identifier: 2019/PID14240).

## Funding

This study was funded by the NSW Health Prevention Research Support Program grant to the Centre for Infectious Diseases and Microbiology-Public Health. R.R. and T.G. are supported by NHMRC Investigator grants (GNT2018222 and GNT2025445)

## Notes

### Competing Interest Statement

The authors have declared no competing interest.

