## Supplementary Figures for "Targeted Culture-Independent Sequencing Identifies Emergence of Macrolide-Resistant *Bordetella Pertussis* in Australia"

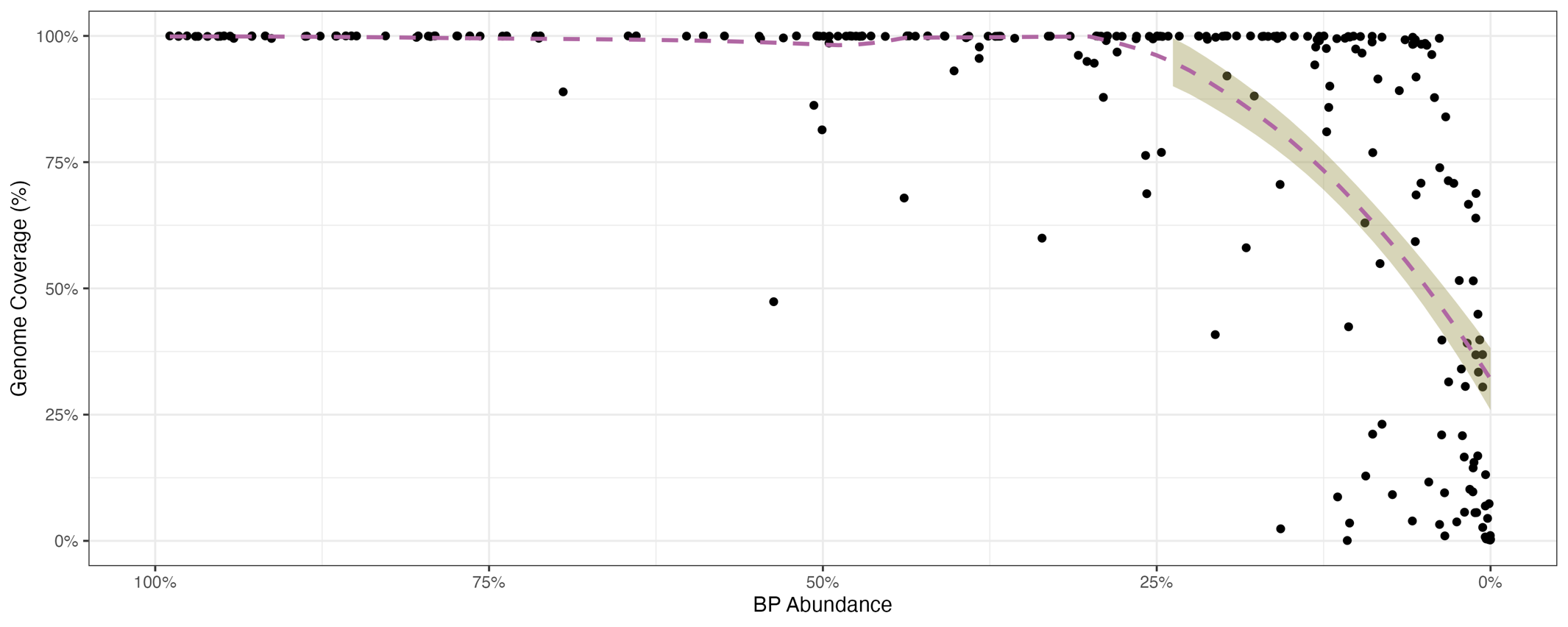

**Figure S1: (A)** Scatterplot of genome coverage and capture efficiency of *Bordetella pertussis* within the clinical specimen. with the purple dashed representing the binomial GLM line, and olive shading representing the 95% confidence interval. Even despite high BP abundance, low genome coverage was still observed.

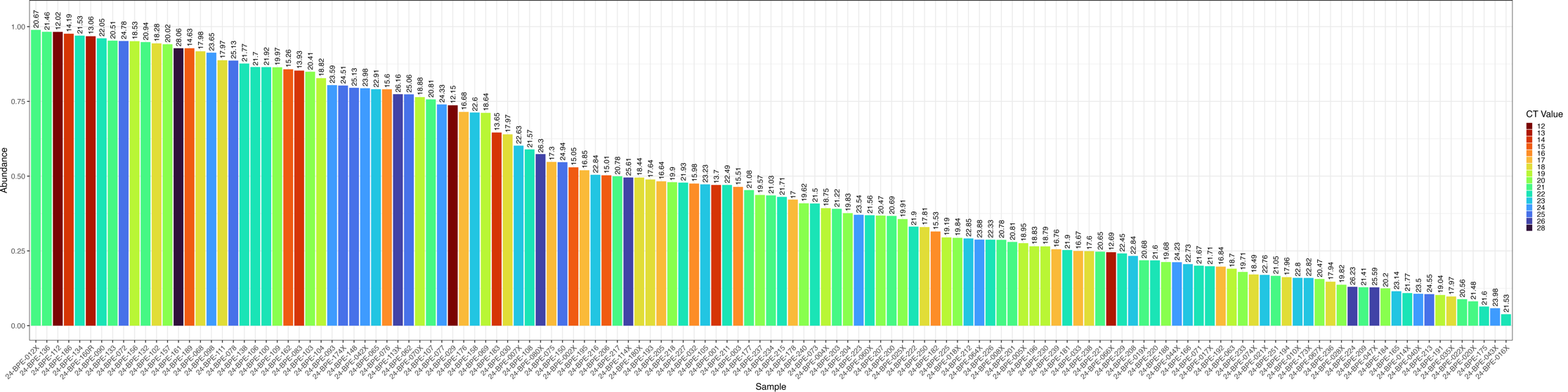

**Figure S2: (A)** Bar chart of *B. pertussis* abundance across successful genomes, demonstrating the importance of *B. pertussis* abundance with the clinical specimen, as well as CT value.

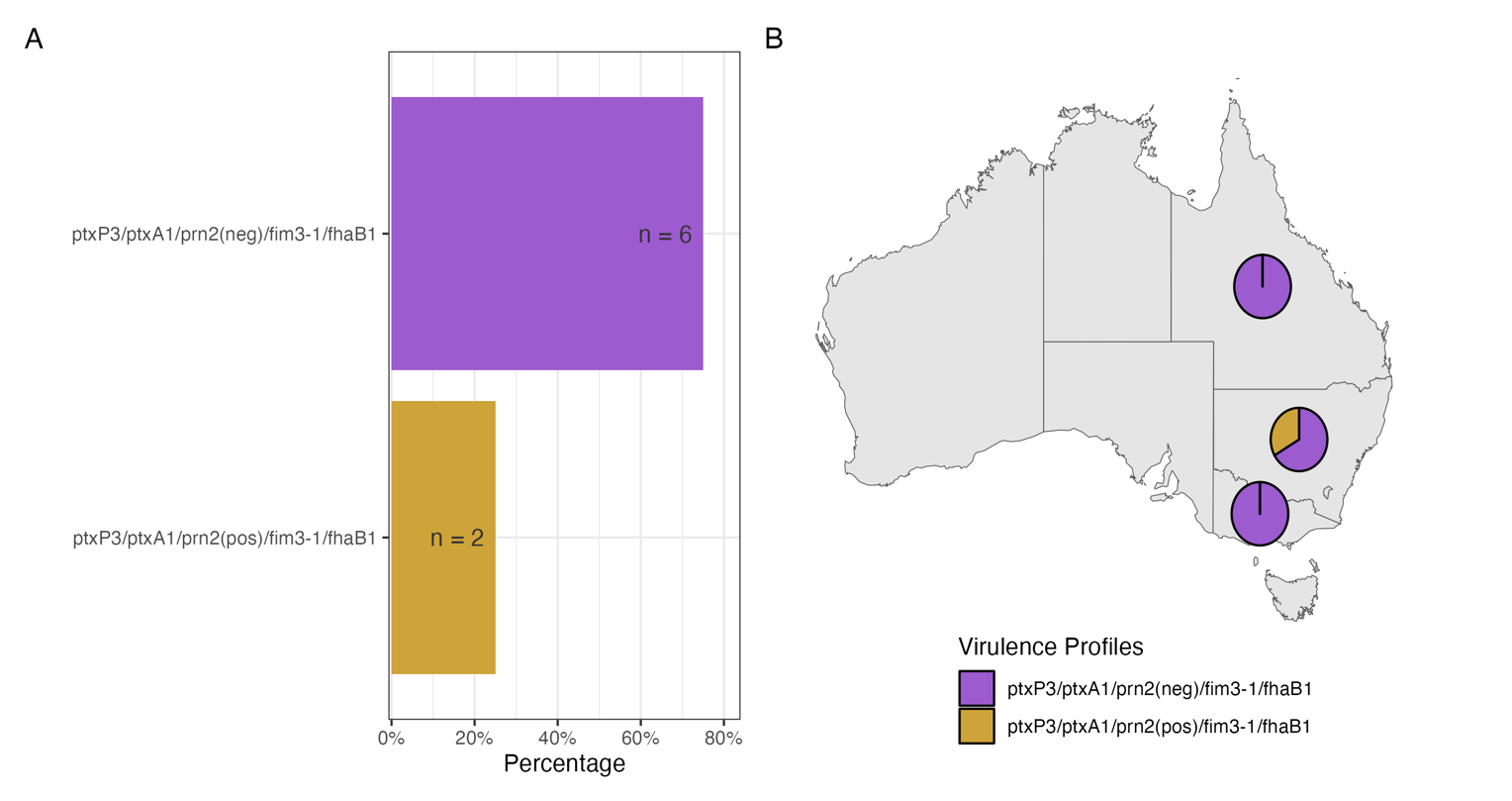

**Figure S3:** Distribution of virulence profiles in MRBP (n = 6) observed during the 2024 B. pertussis outbreak. Colours are coordinated to colours utilised in Figure 1.
